# Synchronous emergence of *Streptococcus pyogenes emm* type 3.93 with unique genomic inversion among invasive infections in the Netherlands and England

**DOI:** 10.1101/2024.06.20.24308992

**Authors:** Matthew A. Davies, Brechje de Gier, Rebecca L Guy, Juliana Coelho, Alje P. van Dam, Robin van Houdt, Sébastien Matamoros, Marit van den Berg, Patrick E. Habermehl, Kartyk Moganeradj, Yan Ryan, Steve Platt, Henry Hearn, Eleanor Blakey, Darren Chooneea, Bart J.M. Vlaminckx, Theresa Lamagni, Nina M. van Sorge

## Abstract

A global increase in the incidence of invasive group A streptococcal (iGAS) infections was observed after lifting COVID-19-related restrictions in 2022 with dominance of M1_UK_ in many countries. After seasonal declines in iGAS incidence during the summer of 2023, simultaneous, rapid expansion of a previously rare *emm* type 3.93 was seen in both England and the Netherlands from November 2023, causing 20% and 60% of all iGAS cases, respectively, within 4 months. *Emm*3.93 was associated with iGAS in children 6-17 years of age and with increased risk of pneumonia/pleural empyema and meningitis in both countries. No significant excess risk of death was identified for *emm*3.93 compared to other types. Genomic analysis of historic and contemporary *emm*3.93 isolates revealed the emergence of three new clades with a previously undescribed genomic inversion. Our findings underscore the value of molecular surveillance, including long-read sequencing, in identifying clinical and public health threats.

## Introduction

In several countries, including the Netherlands and England, an increase in invasive group A streptococcal (iGAS) infections was observed after lifting the COVID-related restrictions in 2022/2023 (1, 2). The reason for this sudden upsurge has yet to be completely explained, but likely resulted from a perfect storm scenario: increased population susceptibility to *S. pyogenes* infections after two years of reduced circulation coinciding with increased incidence of predisposing viral infections and further expansion of the *emm*1.0 sublineage M1UK (3), which produces increased amounts of the virulence factor and superantigen SpeA compared to its progenitor M1_global_ (3). Similar to many other countries, *emm*1.0 was the dominant *emm* type in the Netherlands and England (1, 4, 5). However, from June 2023 onward, the proportion of *emm*1.0 among clinical *S. pyogenes* isolates diminished in both countries, coinciding with a seasonal decrease in iGAS incidence. Subsequently, an increase in a previously rare type, *emm*3.93, was identified among iGAS patients in both countries from November 2023 onward. We report a cross-border increase in iGAS with increased infections in particular age groups and specific clinical manifestations related to a unique *emm*3.93 variant.

## Methods

### S. pyogenes isolate collection, emm typing, and genome sequencing

Since April 2022, all Medical Microbiology laboratories in the Netherlands are requested to submit *S. pyogenes* isolates from patients with iGAS disease (defined as detection of *S. pyogenes* at a normally sterile-site or from a non-sterile-site in combination with a clinical manifestation of iGAS) to the Netherlands Reference Laboratory for Bacterial Meningitis (NRLBM, Amsterdam UMC, Amsterdam) for *emm* typing as part of national bacteriological surveillance. Isolates are submitted with limited patient information (patient birth year, sex, postcode). In addition to recent isolates from prospective bacteriological surveillance, an existing bacterial isolate collection (5), consisting of *S. pyogenes* carriage isolates (2009-2023; n=349) and iGAS disease (2009-2019; n=272) was screened for prevalence of *emm*3.93. In England, Medical Microbiology laboratories are requested to submit *S. pyogenes* isolates to the UK Health Security Agency (UKHSA, London, UK) reference laboratory for *emm* typing as part of active surveillance.

*Emm*-genotyping was performed by conventional PCR amplification and subsequent Sanger sequencing of the 180-bp hypervariable domain of the *emm* gene according to the CDC protocol (6). Short-read sequence (Illumina) data was available or generated for 104 *emm*3.93 isolates, including 11 strains from the Netherlands, 9 from New Zealand and 84 from England. Additionally, long-read (Oxford Nanopore) sequencing was performed for 14 of these *emm*3.93 isolates, five from the Netherlands and nine from England, for hybrid assembly of the complete genome. Raw sequencing data and complete genomes are available from NCBI under the accession numbers in Supplementary File 1.

### Genome assembly

Raw Illumina reads were processed with Trimmomatic v0.39 (7) to remove adaptor sequences and bases of insufficient quality with parameters ILLUMINACLIP:TruSeq3-SE:2:30:10 LEADING:3 TRAILING:3 SLIDINGWINDOW:4:15 MINLEN:36. Trimmed reads were *de novo* assembled into scaffolds with SPAdes v3.15.5 (8) and assembly quality was assessed with QUAST v5.0.2 (9). Raw Nanopore reads were processed with Porechop v0.2.4 (10) with default parameters, assembled with Flye v2.9.2 (11) with parameters -g 5.6m and -i 2. Closed genomes were created with hybrid assembly via Unicycler v0.4.8 (12) with default parameters.

### Detection of genomic rearrangements

To determine the presence of the genomic inversion, complete genomes were annotated with bakta (13) then aligned and visualized with Easyfig (14) and its default use of blastN (15). Location of prophage sequences Φ315.1 (NCBI accession: NC_004584.1) and Φ315.2 (NCBI accession: NC_004585.1) were determined with blastN with default parameters. Whole genome alignment dotplots were produced with the web interface version of D-GENIES (16) using built-in aligner Mashmap v2.0.

### Core SNP phylogenetic analysis

A core SNP alignment was created by calling SNPs with snippy v4.6.0 (https://github.com/tseemann/snippy) against reference genome MGAS315 (NCBI accession: NC_004070.1) with default parameters. Gubbins v2.3.4 (17) was used to detect and mask sites of recombination and a phylogenetic inference by maximum likelihood was performed on the subsequent alignment by IQ-TREE (18) using the substitution model ‘GTR +G +I’, and visualised in iTOL v6.9 (19). The lineage-defining SNPs were determined to be synonymous, non-synonymous or insertion/deletion by snippy. The SNP distance matrix was created with snp-dists v0.8.2 (https://github.com/tseemann/snp-dists) and the hierarchical clustering and visualization was performed by R (20) package pheatmap.

### Clinical information of iGAS patients

For the Netherlands, clinical manifestations of iGAS with *S. pyogenes* cultured from any site are notifiable by law since January 2023. Pseudonymized notifications, which contain information on the disease presentation, onset and when available *emm* type, are analyzed by the National Institute for Public Health and the Environment (RIVM, Bilthoven, the Netherlands). Unknown *emm* types are supplemented by probabilistic linkage of notifications to NRLBM data, on birth year, gender, postcode and date of diagnosis.

In England, iGAS is a notifiable disease since April 2010 (21). Notifications include date of birth, sex, and postcode, and date of specimen. Clinical presentation is not collected, but focus of infection was inferred from specimen type, in particular, cerebrospinal fluid (CSF), pleural fluid and synovial fluid/joint specimens. All iGAS notifications were submitted to the NHS Demographic Batch Tracing Service to identify date of death (22). All-cause mortality was calculated using all deaths within 7 days of positive iGAS specimen, including all post-mortem diagnoses. Multivariable logistic regression was used to assess all-cause mortality and to compare *emm*3.93 with other contemporaneous *emm* types. Antimicrobial susceptibility testing was identified from routine laboratory surveillance data.

### Ethics statement

All studies were conducted in accordance with the European Statements for Good Clinical Practice and the Declaration of Helsinki of the World Medical Association. For the Netherlands, invasive *S. pyogenes* isolates were collected as part of routine care. This study was not reviewed by an ethical review board, as it is based on anonymized surveillance data. UKHSA have legal permission under Regulation 3 of the Health Service (Control of Patient Information) Regulations 2002 to process patient identifiable information without consent.

This process considers the ethics and purpose of collecting and analyzing the data, and as such ethical approval was not separately sought for this work.

## Results

### Emm3.93 epidemiology among iGAS patients

A total of 798 and 1,510 iGAS notifications with a disease onset date between 1 November 2023 and 31 March 2024 were extracted from the notification databases of the Netherlands (on 11 April 2024) and England (on 12 April 2024), respectively. Of the Dutch and English cases, respectively 665 (83.3%) and 1,351 (89.5%) isolates had a known *emm* type. In both countries, the monthly number of submitted *S. pyogenes* isolates from iGAS patients showed a regular seasonal decrease between June and October 2023, coinciding with a decrease in the proportion of *emm*1.0 (Figure 1). From November 2023 onward, iGAS cases increased in line with usual seasonal patterns (Figure 1). Noticeably, the proportion of *emm*3.93 among typed iGAS isolates in the Netherlands increased from 8% (6/73) in November 2023, peaking at 61% (126/207) in February 2024, and from 4% (8/219) in November 2023 to 24% (66/274) in March 2024 in England (Figure 1). From available historic Dutch *S. pyogenes* isolates from iGAS patients and asymptomatic carriers (2009–2019), *emm*3.93 was rare, with only 6 out-of-577 (1%) typed strains. Similarly, *emm*3.93 only constituted 3% (400/11,194) of *S. pyogenes* isolates in England between 2016-2019. A short upsurge in prevalence to 9% (224/2,597) was seen in 2018, but afterwards *emm*3.93 strains decreased to <1% typed isolates per year (23). Phenotypic antimicrobial susceptibility testing demonstrated low resistance to clindamycin (1.4% (CI 0.0-7.4%)), erythromycin (1.4% (CI 0.0-7.5%)), or tetracycline (0% (CI 0-4.4%)) in *emm*3.93 specimens in England between November 2023 and March 2024.

**Figure 1.**
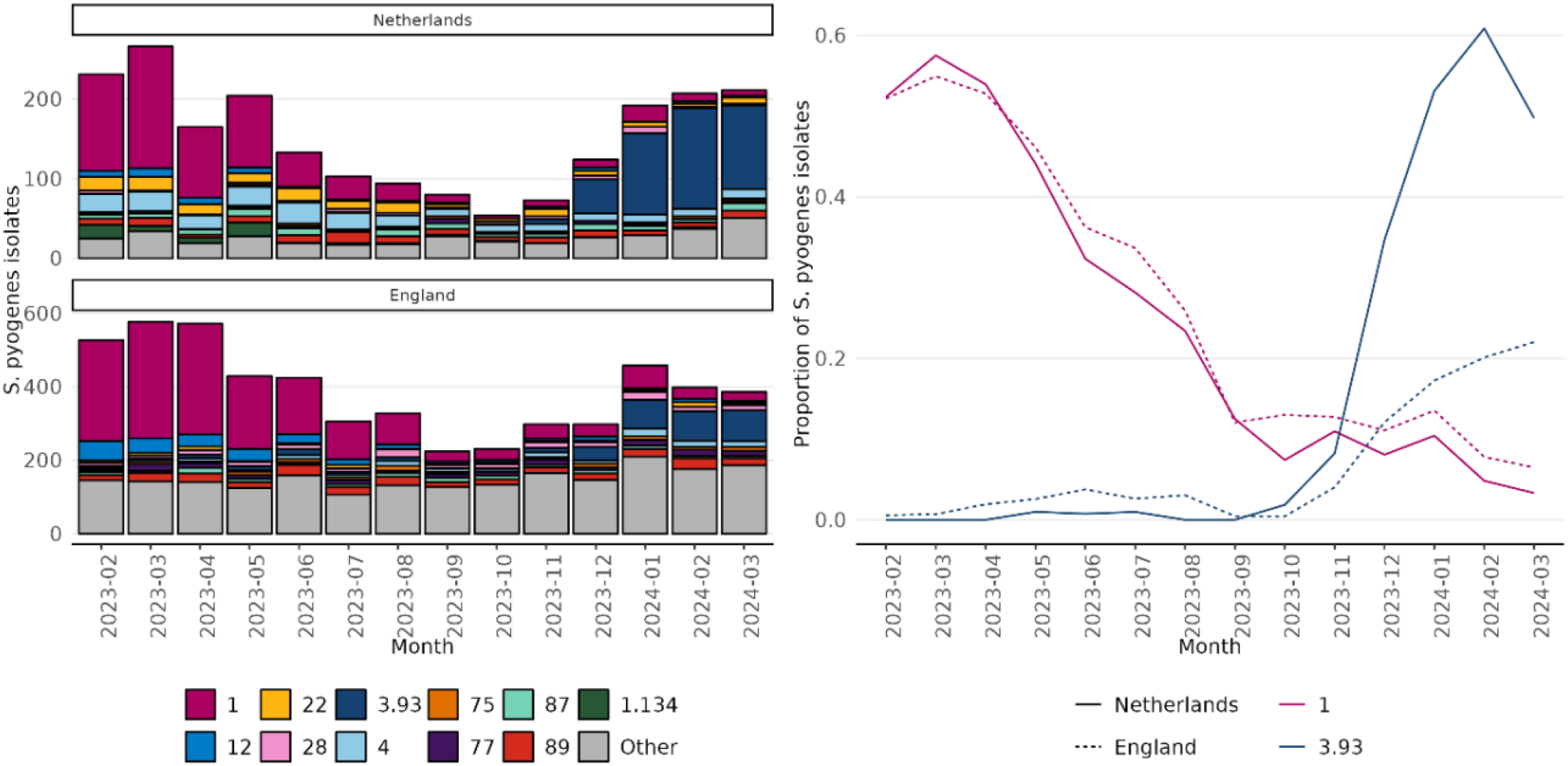
Absolute number of invasive *S. pyogenes* isolates per *emm* type and month (A) and proportion of *emm* types 1.0 and 3.93 among all typed invasive *S. pyogenes* isolates (B), 1 February 2023 – 31 March 2024 in the Netherlands and England.

### Clinical presentation and case-fatality rate of emm3.93 versus other emm types

A multivariable logistic regression model with covariates age group, sex, and month of disease onset revealed that children aged 6-17 years old had higher risk of iGAS caused by *emm*3.93 compared to other *emm* types in both countries (Table 1). In addition, in England, *emm*3.93 also affected children 0-5 years of age significantly more often (aOR 1.7, 95% CI 1.1-2.8; Table 1). Conversely, *emm*3.93 was recovered significantly less often from patients aged 18-44 years old in both countries (Table 1). There was a slight association of *emm*3.93 with female sex compared to other types (aOR 1.4, 95% CI 1.0-1.8) in England, which was not observed in the Netherlands (Table 1).

**Table 1.**
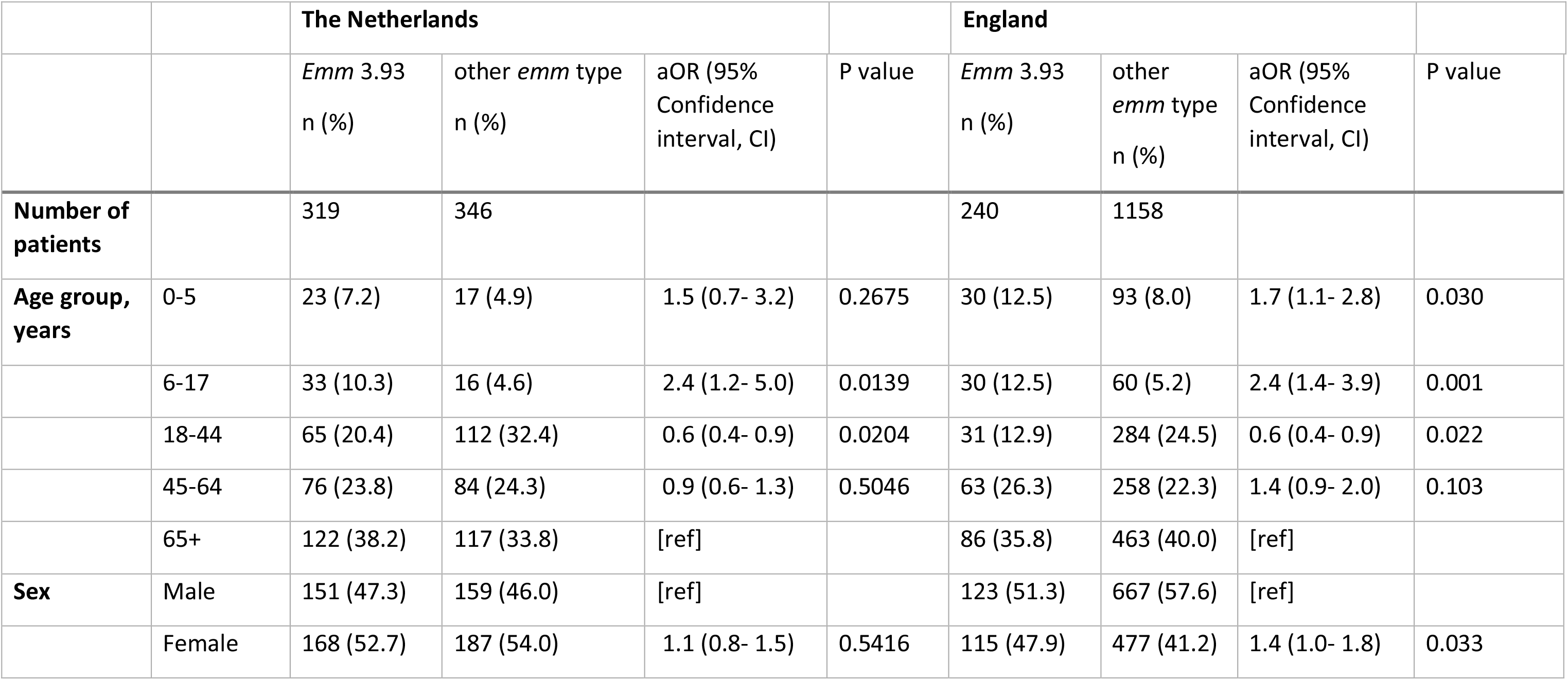
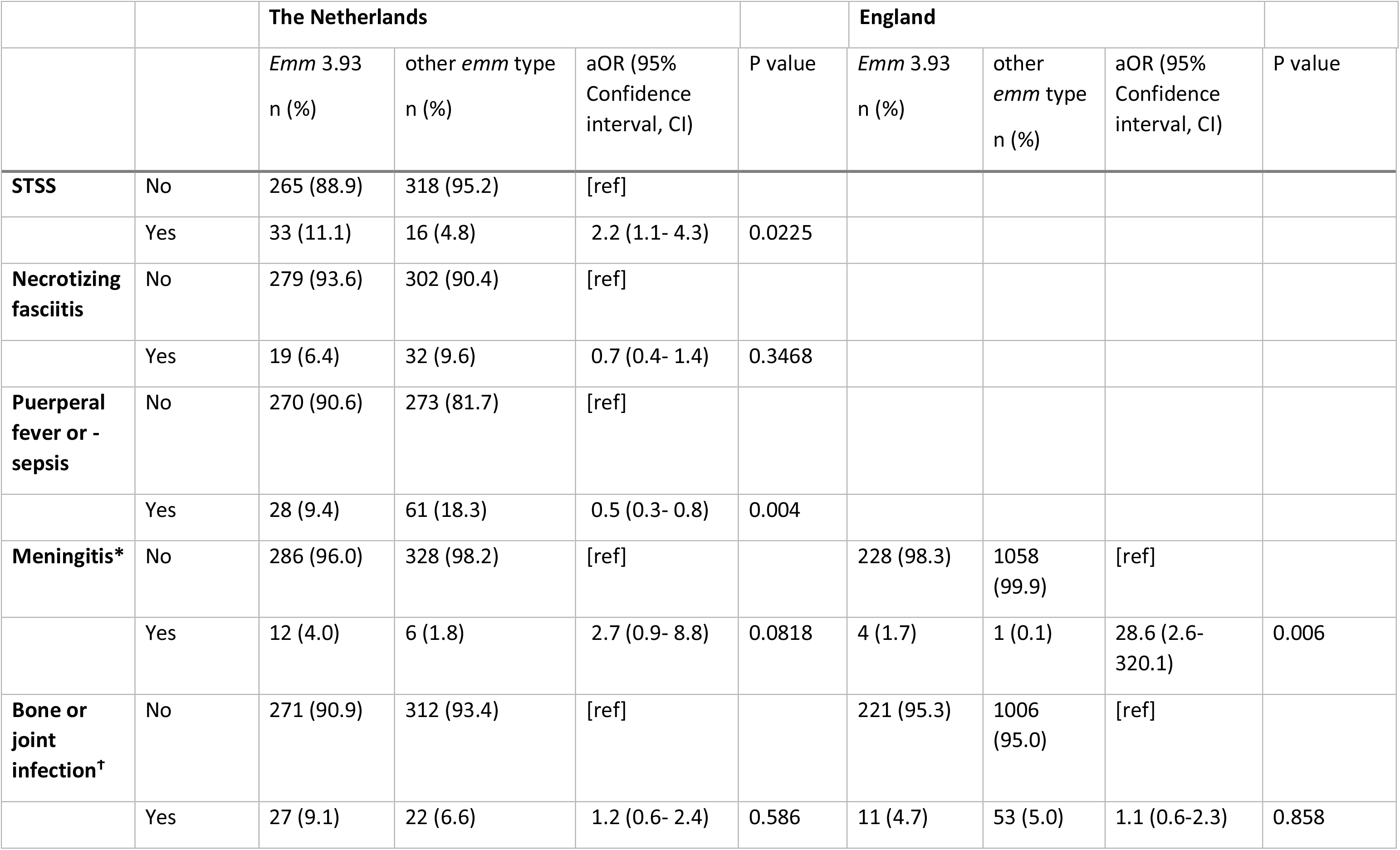
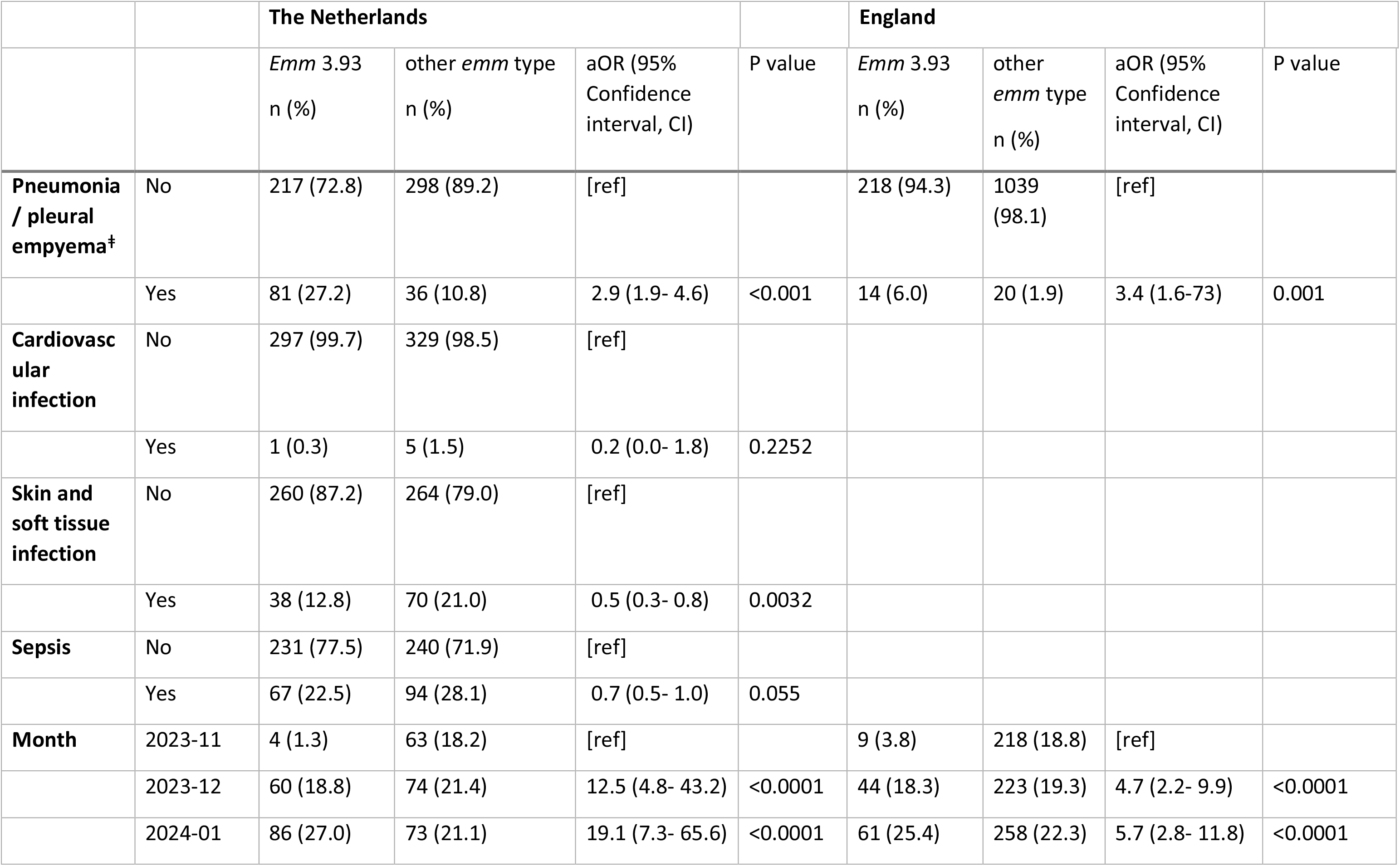
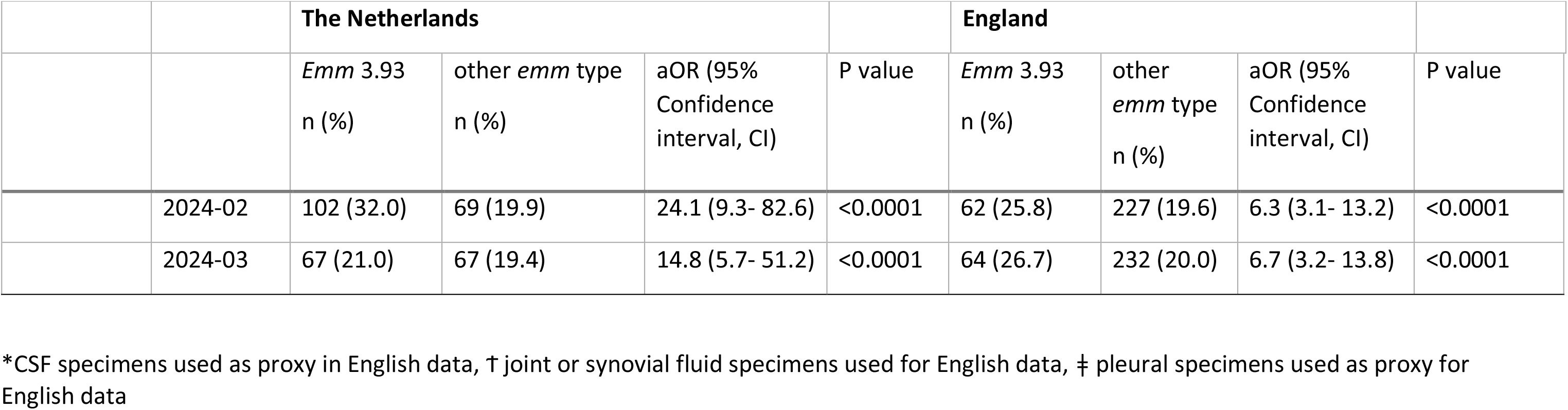
Characteristics of patients with iGAS infection in the Netherlands and England caused by *emm*3.93 or other *emm* types between 1 November 2023 and 31 March 2024. Odds ratios for age group, sex and month are derived from a multivariable model containing these three variables. Odds ratios for clinical presentations are adjusted for age group, sex and month, but the puerperal fever or sepsis aOR is adjusted for month only.

With regard to clinical presentation, *emm*3.93 was significantly associated with pneumonia or pleural empyema in both countries and with STSS (aOR 2.2, 95% CI 1.1-4.3) in the Netherlands (Table 1). Furthermore, *emm*3.93 showed an increased risk for meningitis (Table 1), which was significant in England (aOR 28.6, 95% CI 2.6-320.1) but not in the Netherlands (aOR 2.7, 95% CI 0.9-8.8). In the Netherlands, *emm*3.93 was less often recovered from patients presenting with skin or soft tissue infections or puerperal iGAS infection (aOR 0.5, 95% CI 0.3-0.8; Table 1).

Seven-day (all-cause) CFR data were available for England only. CFR for *emm*3.93 was 13.8% (32/232, 95% CI 9.6-18.9%) compared to 12.0% (127/1059, 95% CI 10.2-14.1%) for other types between November 2023 and March 2024. A multivariable logistic regression adjusting for age, sex and specimen month and *emm* type (*emm*3.93 *vs* other) did not identify a significant difference for seven-day mortality between types (aOR 1.2, 95% CI 0.8-1.8), however significant differences were noted by age group and sex (Table 2).

**Table 2.** Characteristics of patients with iGAS infection caused by *emm*3.93 or other *emm* types according to whether death (all-cause) was recorded within 7 days of diagnosis, England, 1 November 2023 and 31 March 2024. Odds ratios for *emm* type, age group, sex and month are derived from a multivariable model containing these four variables.

|  |  | Died within 7 days<br>n (%) | Alive at 7 days<br>n (%) | Adjusted odds ratio (OR)<br>(95% Confidence interval, CI) | P value |
| --- | --- | --- | --- | --- | --- |
|  |  | 159 | 1132 |  |  |
| **Emm type** | other | 127 (12.0) | 932 (88.0) | [ref] |  |
|  | *emm3.93* | 32 (13.8) | 200 (86.2) | 1.2 (0.8-1.8) | 0.501 |
| **Age group,**<br>**years** | 0-17 | 11 (5.7) | 182 (94.3) | 0.3 (0.1-0.5) | <0.0001 |
|  | 18-44 | 10 (3.5) | 276 (96.5) | 0.2 (0.1-0.3) | <0.0001 |
|  | 45-64 | 42 (14.4) | 250 (85.6) | 0.7 (0.5-1.1) | 0.146 |
|  | 65+ | 96 (18.5) | 424 (82.5) | [ref] |  |
| **Sex** | Male | 77 (10.6) | 651 (89.4) | [ref] |  |
|  | Female | 82 (14.6) | 481 (85.4) | 1.4 (1.0-2.0) | 0.045 |
| Month | 2023-11 | 24 (11.4) | 187 (88.6) | [ref] |  |
|  | 2023-12 | 32 (12.8) | 218 (87.2) | 1.1 (0.6-2.0) | 0.663 |
|  | 2024-01 | 44 (14.9) | 251 (85.1) | 1.3 (0.8-2.3) | 0.339 |
|  | 2024-02 | 34 (12.6) | 237 (87.4) | 1.2 (0.7-2.1) | 0.583 |
|  | 2024-03 | 25 (9.5) | 239 (90.5) | 0.8 (0.4-1.5) | 0.503 |

### Genomic analysis of emm3.93 reveals a unique genomic inversion associated with surge isolates

Phylogenetic analysis of the core SNP genomes of 104 *emm*3.93 from the Netherlands, England, and New Zealand showed that recently expanded *emm*3.93 isolates clustered into three distinct clades (Figure 2A). SNP distances indicated that within-clade variation was <10 core SNPs and between-clade SNP distance was typically 20-50 SNPs (Supplementary Figure S1). Lineage-defining SNPs are described in Supplementary Table 1.

**Figure 2.**
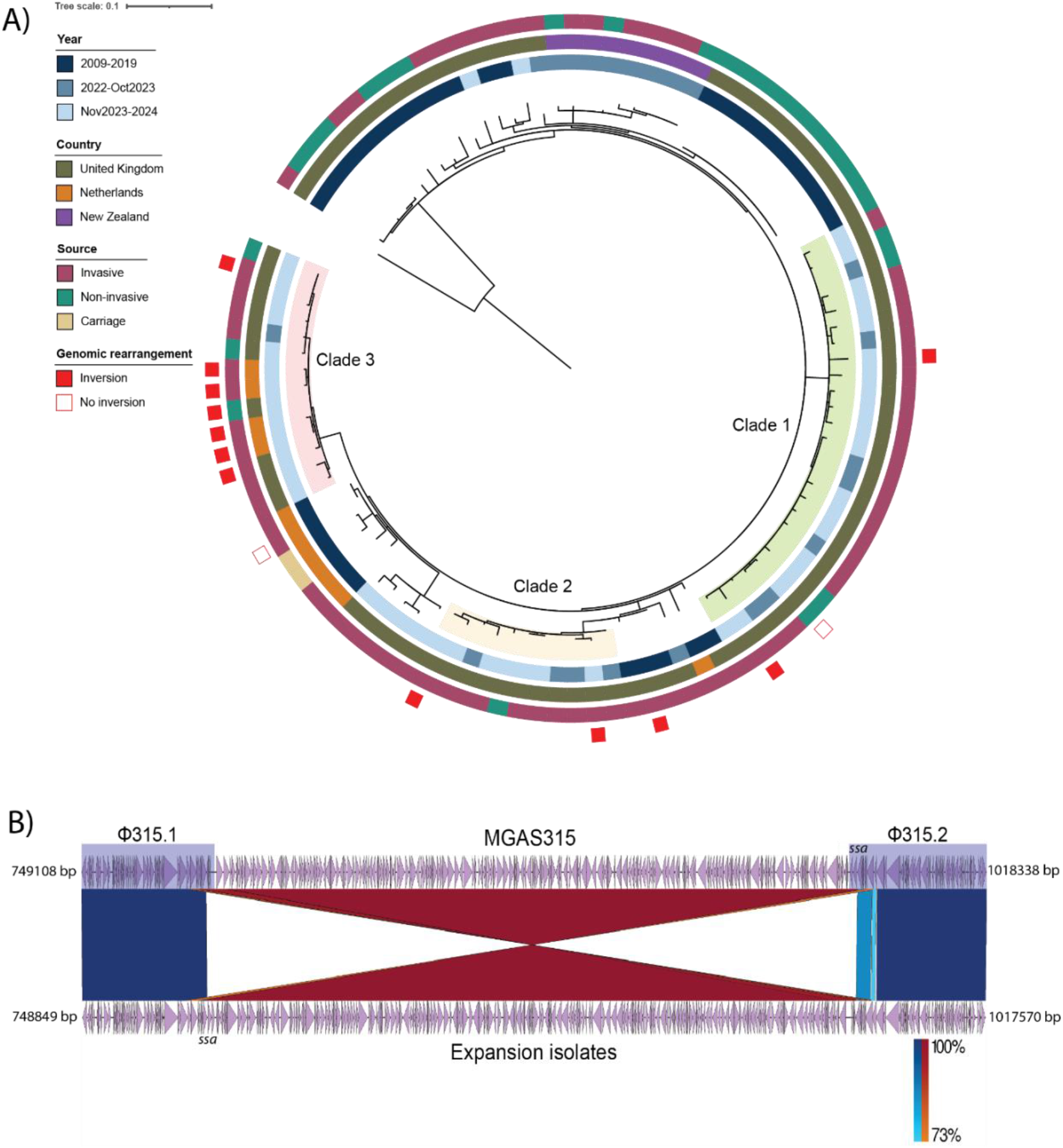
(A) Maximum-likelihood phylogenetic tree from the core genome SNP alignment of 104 *emm*3.93 isolates from the Netherlands, England and New Zealand (2009–2024), against MGAS315 reference genome (NC_004070). Tree was rooted on MGAS315 and colored rings outside the tree represent year of isolation, country, source and presence of genomic inversion represented in (B). (B) Recombination event between MGAS315 reference genome (top) and a Clade 3 isolate (bottom) representative for all isolates marked as containing an inversion. Prophage regions Φ315.1 and Φ315.2 are highlighted in MGAS315 and purple arrowheads indicate genes predicted with bakta, with superantigen gene *ssa* highlighted.

Alignment of closed genomes from hybrid assemblies of four Dutch and three English upsurge iGAS isolates from clade 3 against reference genome MGAS315 revealed a ∼200kb genomic inversion around the terminus and between prophages Φ315.1 and Φ315.2 in the recently expanded isolates (Figure 2B). Interestingly, the phage-encoded superantigen gene *ssa* is located at the internal boundary of the genomic inversion, moving ∼200 kb back in the genome and crossing the *ter* site. Compared to other publicly-available closed *emm*3.1 and *emm*3.93 genomes, the genomic rearrangement was unique (Supplementary Figure S2). A similar genome inversion was present in four English iGAS isolates from the other *emm*3.93 surge clades 1 and 2 (Figure 2A), but not in a Dutch *emm*3.93 isolate from 2017 that is phylogenetically close to clade 3 and one *S. pyogenes* isolate in clade 1 from a patient with non-invasive disease (Figure 2A). Interestingly, one of the invasive *emm*3.93 isolates isolated in England in 2017 also showed the unique genomic inversion (Figure 2A), which may indicate that this inverted *emm*3.93 variant was already circulating before the current upsurge in 2023/2024.

## Discussion

We report the rapid synchronous upsurge of *S. pyogenes emm* type 3.93 in the Netherlands and England between November 2023 and March 2024. Epidemiological analysis showed that *emm*3.93 was isolated significantly more often in children with iGAS aged 6-17 years old in both countries. The Dutch data revealed a significant association between *emm*3.93 and pneumonia or pleural empyema, which was corroborated by an increase of *emm*3.93 among pleural isolates in England. Although the numbers were low, *emm*3.93 showed an increased risk for causing meningitis, which reached significance for the English data.

Furthermore, *emm*3.93 was associated with STSS in the Netherlands. Despite these concerning associations, the English data showed no significant elevation in 7-day mortality for *emm*3.93 compared to mortality associated to other *emm* types.

Genomic analysis revealed three distinct clades among recent *emm*3.93 surge isolates, and analysis of complete genomes revealed the presence of a ∼200kb genomic inversion in all but one recent isolate from these clades. This isolate without the inversion belonged to clade 1 and was collected from a non-invasive wound swab. In addition, a Dutch invasive *emm*3.93 isolate isolated in 2017 had the same genome arrangement as the reference isolate MGAS315, i.e. lacking the genomic inversion of clade 3 isolates, and differed only an average of approximately 20 core SNPs. The relatively low number of point mutations compared to clade 3, and the sharing of a closest common ancestor with two carriage isolates, suggests that the genomic inversion could explain the specific epidemiological characteristics of iGAS-associated *emm*3.93 strains. However, an invasive English *emm*3.93 isolate from 2017 also had the genomic inversion. Interestingly, *emm*3.93 caused a small surge of iGAS in England in 2018 but did not expand to the levels observed in 2023-2024 nor did other countries report on iGAS caused by *emm*3.93. Perhaps this isolate was a precursor to the 2023-2024 surge but did not result into a significant advantage and expansion in the pre-COVID-19 climate as a result of sufficient high levels of population immunity.

Inverted repeats between prophages Φ315.1 and Φ315.2 have likely permitted the observed prophage-mediated homologous recombination. This phenomenon has been observed before in *emm*3.1 strains, which contained an alternative large-scale genomic rearrangement of 1Mb due to chromosomal inverted repeats and was associated with invasive disease (24). The fact that the phage-encoded superantigen gene *ssa* has shifted location, could affect the transcriptional regulation of *ssa*, which was previously shown to be pyrogenic and toxic in rabbits (25). Additional research is required to demonstrate whether Ssa expression is indeed affected as a result of the observed genomic inversion.

A limitation of the study is the fact that data collected in Public Health registration systems differ between England and the Netherlands. Differences exists between clinical data collected and the Netherlands lacks a formal registration for mortality after infection. Another limitation is the low number of genomes sequenced Dutch isolates collected during the recent outbreak. Whereas all four recent Dutch 3.93 isolates are located in a single cluster, it may be that a significant number of other so far non-sequenced isolates would fall into the other outbreak-associated clusters.

Combined epidemiological and molecular surveillance is key to detect emergence of new -more virulent-variants and to rapidly assess their clinical and epidemiological relevance. *Emm* typing revealed a replacement of *emm*1.0 by *emm*3.93 as dominant type from November 2023 onwards, continuing over a few months. It is possible that population immunity to *emm*1.0 developed during 2023, leaving increased susceptibility to potentially new virulence traits of the new clades of *emm*3.93. Alternatively, the genomic inversion may have resulted in a competitive advantage of *emm*3.93 over *emm*1.0. In the context of the current upsurge of *S. pyogenes emm*3.93, international collaboration proved invaluable to assess the spread of the new clades and a more comprehensive picture of the associations of *emm*3.93 with both clinical manifestations and case fatality. Additionally, a combination of long- and short read sequencing was invaluable to reveal the extent of genetic diversity for one of the emerged *emm*3.93 clades.

## Data Availability

The data underlying this article cannot be shared publicly due to patient confidentiality and deductive disclosure issues. Aggregated data will be shared on reasonable request to the corresponding author. Raw sequencing data and complete genomes are available from NCBI under the accession numbers listed in Supplementary File 1.

## Acknowledgements

We acknowledge our colleagues of the Institute of Environmental Science and Research (ESR) in New Zealand, in particular Dr. Una Ren, for sharing genome sequences of recent *emm*3.93 strains. We would also like to thank all microbiology laboratories for submitting isolates to the UKHSA national reference laboratory and the NRLBM. This report would not have been possible without the contribution of clinical and laboratory colleagues across the healthcare systems in England and the Netherlands.

## Data sharing statement

The data underlying this article cannot be shared publicly due to patient confidentiality and deductive disclosure issues. Aggregated data will be shared on reasonable request to the corresponding author.

