## Supplementary information for "Synchronous emergence of *Streptococcus pyogenes emm* type 3.93 with unique genomic inversion among invasive infections in the Netherlands and England"

**Supplementary Table 1:**

Lineage-specific single nucleotide polymorphisms (SNPs). Clade-specific SNP annotations against reference strain (Ref) MGAS315 (NC_004070.1), determined with Snippy. S/NS refers to synonymous and non-synonymous mutations, respectively.

| Clade | MGAS315 Location | Locus tag | Gene | Product | S/NS | Amino acid change | Ref | SNP/indel |
|---|---|---|---|---|---|---|---|---|
| **Clade 1 (26 isolates)** | 298891 | SPYM3_RS01565 | yidC | membrane protein insertase YidC | NS | Arg305Gln | C | T |
| | 347532 | Intergenic | | | Intergenic | | CA | C |
| | 401009 | SPYM3_RS02125 | ccpA | catabolite control protein A | NS | Ala167Asp | C | A |
| | 433606 | SPYM3_RS10295 | | DUF2326 domain-containing protein | Frameshift | Lys158fs | AT | A |
| | 529296 | Intergenic | | | Intergenic | | A | G |
| | 591410 | Intergenic | | | Intergenic | | AG | A |
| | 700812 | SPYM3_RS03535 | recJ | single-stranded-DNA-specific exonuclease RecJ | NS | Met315Val | A | G |
| | 799621 | SPYM3_RS04090 | | response regulator transcription factor | NS | Gly82Ser | G | A |
| | 810674 | SPYM3_RS04135 | pbp3 | D-alanyl-D-alanine carboxypeptidase PBP3 | NS | Thr265Ser | C | G |

| | | | | | | | | |
|---|---|---|---|---|---|---|---|---|
| | 947767 | SPYM3_RS04805 | | restriction endonuclease subunit S | S | Gly48Gly | G | C |
| | 1068134 | SPYM3_RS05460 | | methionyl aminopeptidase | S | Leu255Leu | T | C |
| | 1217111 | Intergenic | | | Intergenic | | T | C |
| | 1434583 | SPYM3_RS07490 | | PBSX family phage terminase large subunit | NS | Phe266Tyr | A | T |
| | 1581156 | SPYM3_RS08200 | | magnesium transporter CorA family protein | S | Leu262Leu | A | G |
| | 1609622 | SPYM3_RS08345 | | MerR family transcriptional regulator | S | Asp116Asp | G | A |
| **Clade 2 (11 isolates)** | 184143 | SPYM3_RS01030 | speG | streptococcal pyrogenic exotoxin SpeG | S | Ser205Ser | T | C |
| | 642839 | SPYM3_RS03245 | ideS | immunoglobulin G-degrading enzyme IdeS | NS | Glu66Lys | C | T |
| | 833981 | Intergenic | | | Intergenic | | T | TA |
| | 1147260 | SPYM3_RS05820 | | hypothetical protein | NS | Thr59Ile | G | A |
| | 1524461 | SPYM3_RS07970 | grpE | nucleotide exchange factor GrpE | NS | Leu103Ile | G | T |
| | 1740613† | SPYM3_RS08960 | | pneumococcal-type histidine triad protein | NS | Ala172Val | G | A |
| | 1763604 | SPYM3_RS09050 | ropB | quorum-sensing system transcriptional regulator RopB/Rgg1 | NS | Asp24His | G | C |

| | | | | | | | | |
|---|---|---|---|---|---|---|---|---|
| **Clade 3 (14 isolates)** | 341961 | SPYM3_RS01800 | metG | methionine--tRNA ligase | S | Pro229Pro | A | C |
| | 370021‡ | SPYM3_RS01945 | msrA | peptide-methionine (S)-S-oxide reductase MsrA | NS | Ala158Thr | G | A |
| | 427400 | SPYM3_RS02235 | | serine kinase | NS | Asp145Gly | A | G |
| | 502485 | SPYM3_RS02620 | | (S)-acetoin forming diacetyl reductase | S | Tyr150Tyr | C | T |
| | 629815 | SPYM3_RS03180 | | cation diffusion facilitator family transporter | NS | Pro200Ser | G | A |
| | 987708 | SPYM3_RS05000 | | phage tail spike protein | S | Arg79Arg | C | T |
| | 987714 | SPYM3_RS05000 | | phage tail spike protein | S | delAGTGinsGGTA | CACT | TACC |
| | 1270371 | SPYM3_RS06610 | | hypothetical protein | S | Asp117Asp | C | T |
| | 1281283 | SPYM3_RS06655 | | shikimate kinase | NS | Asn171Lys | G | C |
| | 1300051 | SPYM3_RS06745 | | ROK family protein | S | Asn105Asn | T | C |
| | 1802116 | SPYM3_RS09230 | | APC family permease | NS | Thr122Ile | C | T |

*Also present in the two most related isolates to the clade

†Missing from one isolate in the clade

‡Missing from two isolates in the clade

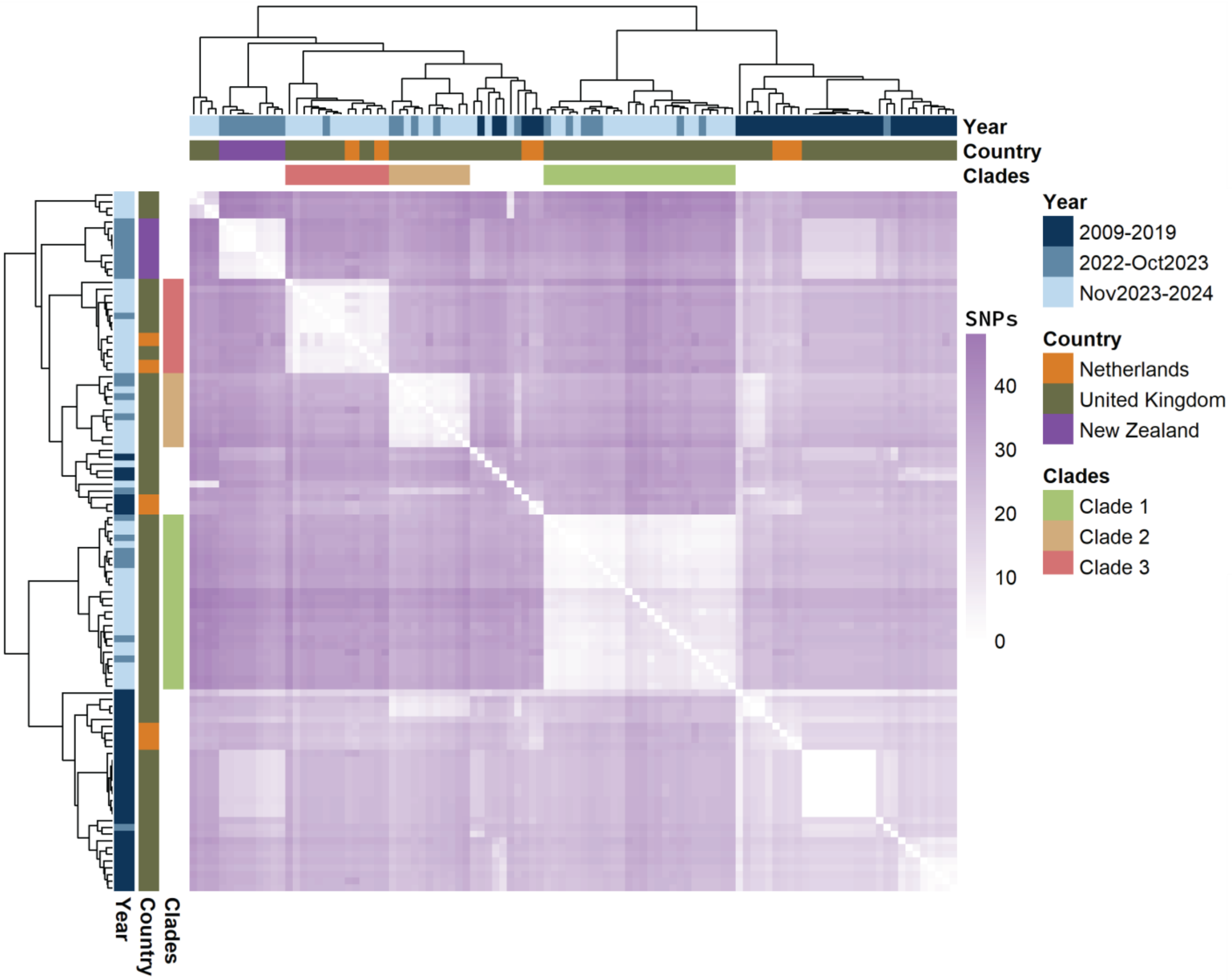

**Supplementary Figure S1.** Core-SNP distance matrix of the core SNP alignment of 104 *emm3.93* genomes from the Netherlands, United Kingdom and New Zealand between 2009-2024. Color in the heatmap indicates the number of SNPs between isolates, with highly similar isolates in white and shifting towards purple as number of SNPs increases. Year, country, and *emm3.93* clades are visualized by both dendrograms, which have been clustered with default hierarchical clustering from R package pheatmap.

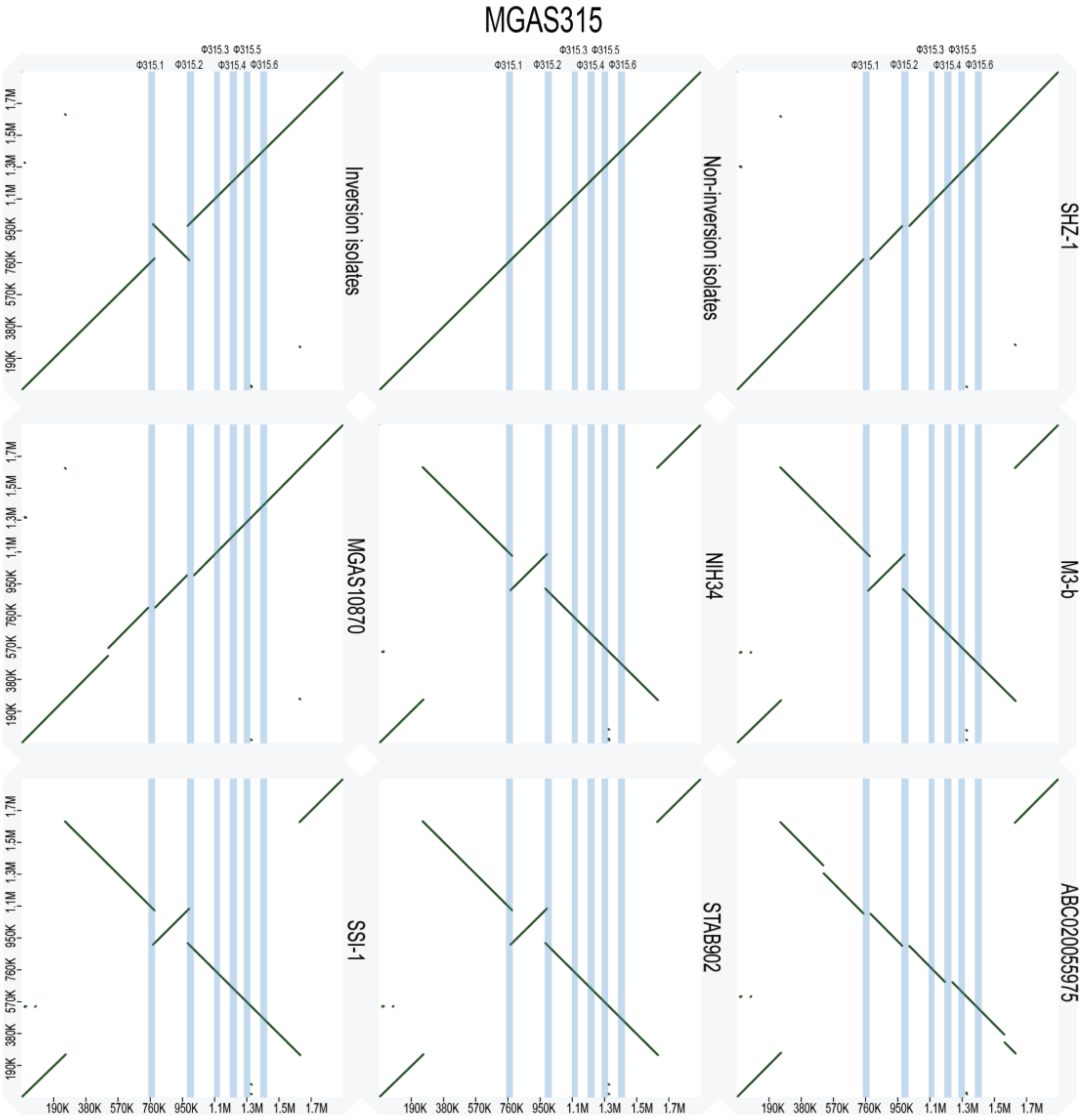

**Supplementary Figure S2.** Dotplots representing pairwise genome alignment of MGAS315 reference genome (NC_004070; x-axes) against either inversion isolates, non-inversion isolates or all publicly available *emm3.1* complete genomes (y-axes). An upwards diagonal black line represents region of shared sequence in the same orientation, whereas a downwards diagonal black line represents region of shared sequence in the opposition orientation. Six known MGAS315 prophages (Φ315.1, Φ315.2, Φ315.3, Φ315.4, Φ315.5, Φ315.6) are visualized as blue vertical lines. White gaps represent no sequence similar shared between the genomes. Plots were generated by D-GENIES.
