## Supplementary File 1 for "Synchronous emergence of *Streptococcus pyogenes emm* type 3.93 with unique genomic inversion among invasive infections in the Netherlands and England"

| Sample ID | Country | Colonisation type | emmtype | collection_year | Collectiondate_period (1=2009-2011; 2=2012-2014; 3=2015-2017; 4=2018-2019; 5=2020-2021; 6=2022-Oct2023; 7=Nov2023-2024) | Source | BioProject | Sample Accession |
|---|---|---|---|---|---|---|---|---|
| 1138712 | Netherlands | Invasive | 3.93 | 2017 | 3 | Blood | PRJNA1125189 | SAMN41885169 |
| 4612210 | Netherlands | Invasive | 3.93 | 2017 | 3 | Blood | PRJNA1125189 | SAMN41885170 |
| 9877795 | Netherlands | Invasive | 3.93 | 2017 | 3 | CSF | PRJNA1125189 | SAMN41885171 |
| 4117681 | Netherlands | Invasive | 3.93 | 2019 | 4 | Blood | PRJNA1125189 | SAMN41885172 |
| 1221089 | Netherlands | Invasive | 3.93 | 2023 | 7 | Blood | PRJNA1125189 | SAMN41885173 |
| 6101976 | Netherlands | Invasive | 3.93 | 2024 | 7 | CSF | PRJNA1125189 | SAMN41885174 |
| 1157077 | Netherlands | Invasive | 3.93 | 2024 | 7 | Sputum | PRJNA1125189 | SAMN41885175 |
| 1111116 | Netherlands | Invasive | 3.93 | 2024 | 7 | CSF | PRJNA1125189 | SAMN41885176 |
| 1001106 | Netherlands | Carriage | 3.93 | 2017 | 3 | Nose | PRJNA1125189 | SAMN41885177 |
| 4610265 | Netherlands | Carriage | 3.93 | 2017 | 3 | Nose | PRJNA1125189 | SAMN41885178 |
| 1211118 | Netherlands | Invasive | 3.93 | 2016 | 3 | Blood | PRJNA1125189 | SAMN41885179 |
| 511605 | England | Invasive | 3.93 | 2017 | 3 | Blood | PRJNA1125189 | SAMN41885180 |
| 511606 | England | Invasive | 3.93 | 2017 | 3 | Blood | PRJNA1125189 | SAMN41885181 |
| 511608 | England | Invasive | 3.93 | 2018 | 4 | Blood | PRJNA1125189 | SAMN41885182 |
| 511614 | England | Invasive | 3.93 | 2018 | 4 | Blood | PRJNA1125189 | SAMN41885183 |
| 511619 | England | Invasive | 3.93 | 2018 | 4 | Blood | PRJNA1125189 | SAMN41885184 |
| 537090 | England | Invasive | 3.93 | 2018 | 4 | Blood | PRJNA1125189 | SAMN41885185 |
| 705173 | England | Invasive | 3.93 | 2018 | 4 | Tissue | PRJNA1125189 | SAMN41885186 |
| 1110171 | England | Invasive | 3.93 | 2023 | 6 | Blood | PRJNA1125189 | SAMN41885187 |
| 1112763 | England | Invasive | 3.93 | 2023 | 6 | Blood | PRJNA1125189 | SAMN41885188 |
| 1112829 | England | Invasive | 3.93 | 2023 | 6 | Blood | PRJNA1125189 | SAMN41885189 |
| 1154616 | England | Invasive | 3.93 | 2023 | 6 | Tissue | PRJNA1125189 | SAMN41885190 |
| 1154654 | England | Invasive | 3.93 | 2023 | 6 | Blood | PRJNA1125189 | SAMN41885191 |
| 1154693 | England | Invasive | 3.93 | 2023 | 6 | Tissue | PRJNA1125189 | SAMN41885192 |
| 1164292 | England | Invasive | 3.93 | 2023 | 6 | Abscess | PRJNA1125189 | SAMN41885193 |
| 1188675 | England | Invasive | 3.93 | 2023 | 6 | Blood | PRJNA1125189 | SAMN41885194 |
| 1193701 | England | Invasive | 3.93 | 2023 | 6 | Pus | PRJNA1125189 | SAMN41885195 |
| 1193716 | England | Non-invasive | 3.93 | 2023 | 6 | Wound swab | PRJNA1125189 | SAMN41885196 |
| 1200347 | England | Invasive | 3.93 | 2023 | 6 | Blood | PRJNA1125189 | SAMN41885197 |
| 1206953 | England | Invasive | 3.93 | 2023 | 6 | Blood | PRJNA1125189 | SAMN41885198 |
| 1219862 | England | Invasive | 3.93 | 2023 | 6 | Blood | PRJNA1125189 | SAMN41885199 |
| 1314957 | England | Invasive | 3.93 | 2023 | 7 | Blood | PRJNA1125189 | SAMN41885200 |
| 1329712 | England | Invasive | 3.93 | 2023 | 7 | Blood | PRJNA1125189 | SAMN41885201 |
| 1339736 | England | Invasive | 3.93 | 2023 | 7 | Blood | PRJNA1125189 | SAMN41885202 |
| 1339738 | England | Invasive | 3.93 | 2023 | 7 | Blood | PRJNA1125189 | SAMN41885203 |
| 1355993 | England | Invasive | 3.93 | 2023 | 7 | Blood | PRJNA1125189 | SAMN41885204 |
| 1355999 | England | Non-invasive | 3.93 | 2023 | 7 | Throat swab | PRJNA1125189 | SAMN41885205 |
| 1356031 | England | Invasive | 3.93 | 2023 | 7 | Blood | PRJNA1125189 | SAMN41885206 |
| 1377859 | England | Invasive | 3.93 | 2023 | 7 | Blood | PRJNA1125189 | SAMN41885207 |
| 1377860 | England | Invasive | 3.93 | 2023 | 7 | Blood | PRJNA1125189 | SAMN41885208 |
| 1380021 | England | Invasive | 3.93 | 2024 | 7 | Blood | PRJNA1125189 | SAMN41885209 |
| 1380060 | England | Invasive | 3.93 | 2024 | 7 | Blood | PRJNA1125189 | SAMN41885210 |
| 1380103 | England | Invasive | 3.93 | 2024 | 7 | Blood | PRJNA1125189 | SAMN41885211 |
| 1389677 | England | Invasive | 3.93 | 2024 | 7 | Blood | PRJNA1125189 | SAMN41885212 |
| 1389695 | England | Invasive | 3.93 | 2024 | 7 | CSF | PRJNA1125189 | SAMN41885213 |
| 1389698 | England | Invasive | 3.93 | 2024 | 7 | Pleural fluid | PRJNA1125189 | SAMN41885214 |
| 1389710 | England | Invasive | 3.93 | 2024 | 7 | Blood | PRJNA1125189 | SAMN41885215 |
| 1389712 | England | Non-invasive | 3.93 | 2024 | 7 | Ear swab | PRJNA1125189 | SAMN41885216 |
| 1389743 | England | Invasive | 3.93 | 2024 | 7 | Tissue | PRJNA1125189 | SAMN41885217 |
| 1397800 | England | Non-invasive | 3.93 | 2024 | 7 | Vaginal swab | PRJNA1125189 | SAMN41885218 |
| 1397823 | England | Non-invasive | 3.93 | 2024 | 7 | Wound swab | PRJNA1125189 | SAMN41885219 |
| 1397858 | England | Invasive | 3.93 | 2024 | 7 | Blood | PRJNA1125189 | SAMN41885220 |
| 1397882 | England | Invasive | 3.93 | 2024 | 7 | Blood | PRJNA1125189 | SAMN41885221 |
| 1413895 | England | Non-invasive | 3.93 | 2024 | 7 | Wound swab | PRJNA1125189 | SAMN41885222 |
| 1414070 | England | Invasive | 3.93 | 2024 | 7 | Blood | PRJNA1125189 | SAMN41885223 |
| 1417878 | England | Invasive | 3.93 | 2024 | 7 | Blood | PRJNA1125189 | SAMN41885224 |
| 1417895 | England | Invasive | 3.93 | 2024 | 7 | Blood | PRJNA1125189 | SAMN41885225 |
| 1420627 | England | Invasive | 3.93 | 2024 | 7 | Blood | PRJNA1125189 | SAMN41885226 |
| 1420644 | England | Invasive | 3.93 | 2024 | 7 | Pleural fluid | PRJNA1125189 | SAMN41885227 |
| 1420649 | England | Non-invasive | 3.93 | 2024 | 7 | Skin swab | PRJNA1125189 | SAMN41885228 |
| 1426265 | England | Invasive | 3.93 | 2024 | 7 | Blood | PRJNA1125189 | SAMN41885229 |
| 1426268 | England | Invasive | 3.93 | 2024 | 7 | Blood | PRJNA1125189 | SAMN41885230 |
| 1426286 | England | Non-invasive | 3.93 | 2024 | 7 | Wound swab | PRJNA1125189 | SAMN41885231 |
| 1426295 | England | Invasive | 3.93 | 2024 | 7 | Blood | PRJNA1125189 | SAMN41885232 |
| 1426298 | England | Invasive | 3.93 | 2024 | 7 | Blood | PRJNA1125189 | SAMN41885233 |
| 1426315 | England | Invasive | 3.93 | 2024 | 7 | Blood | PRJNA1125189 | SAMN41885234 |
| 1427257 | England | Invasive | 3.93 | 2024 | 7 | Aspirate | PRJNA1125189 | SAMN41885235 |
| 1433694 | England | Invasive | 3.93 | 2024 | 7 | Blood | PRJNA1125189 | SAMN41885236 |
| 1441513 | England | Invasive | 3.93 | 2024 | 7 | Blood | PRJNA1125189 | SAMN41885237 |
| 1441515 | England | Invasive | 3.93 | 2024 | 7 | Blood | PRJNA1125189 | SAMN41885238 |
| 1441543 | England | Invasive | 3.93 | 2024 | 7 | Blood | PRJNA1125189 | SAMN41885239 |
| 1441558 | England | Invasive | 3.93 | 2024 | 7 | Blood | PRJNA1125189 | SAMN41885240 |
| 1441578 | England | Invasive | 3.93 | 2024 | 7 | Post mortem swab | PRJNA1125189 | SAMN41885241 |
| 1822122 | England | Invasive | 3.93 | 2023 | 6 | Blood | PRJNA1125189 | SAMN41885242 |
| 1904308 | England | Invasive | 3.93 | 2018 | 4 | Pleural fluid | PRJNA1125189 | SAMN41885243 |
| C_CC09_C_007 | England | Non-invasive | 3.93 | 2019 | 4 | Cough plate | PRJEB43915 | SAMEA8426085 |
| C_CC09_H_010 | England | Non-invasive | 3.93 | 2019 | 4 | Hand swab | PRJEB43915 | SAMEA8426086 |
| C_CC09_T_007 | England | Non-invasive | 3.93 | 2019 | 4 | Throat swab | PRJEB43915 | SAMEA8426087 |
| C_CC38_T_010 | England | Non-invasive | 3.93 | 2019 | 4 | Throat swab | PRJEB43915 | SAMEA8426108 |
| C_CC42_T_010 | England | Non-invasive | 3.93 | 2019 | 4 | Throat swab | PRJEB43915 | SAMEA8426116 |
| C_E1_S_010 | England | Non-invasive | 3.93 | 2019 | 4 | Settle plate | PRJEB43915 | SAMEA8426124 |
| C_E3_S_001 | England | Non-invasive | 3.93 | 2019 | 4 | Settle plate | PRJEB43915 | SAMEA8426125 |

| | | | | | | | | |
|---|---|---|---|---|---|---|---|---|
| C_E4_S_007 | England | Non-invasive | 3.93 | 2019 | 4 | Settle plate | PRJEB43915 | SAMEA8426126 |
| C_E4_S_010 | England | Non-invasive | 3.93 | 2019 | 4 | Settle plate | PRJEB43915 | SAMEA8426127 |
| C_H2_T_010 | England | Non-invasive | 3.93 | 2019 | 4 | Throat swab | PRJEB43915 | SAMEA8426128 |
| ERR1359363 | England | Non-invasive | 3.93 | 2014 | 2 | | PRJEB13551 | SAMEA3930736 |
| ERR1359409 | England | Invasive | 3.93 | 2014 | 2 | | PRJEB13551 | SAMEA3930992 |
| ERR1359410 | England | Invasive | 3.93 | 2014 | 2 | | PRJEB13551 | SAMEA3931030 |
| ERR1359485 | England | Non-invasive | 3.93 | 2014 | 2 | | PRJEB13551 | SAMEA3930732 |
| ERR1359601 | England | Non-invasive | 3.93 | 2014 | 2 | | PRJEB13551 | SAMEA3930878 |
| ERR1359650 | England | Invasive | 3.93 | 2014 | 2 | | PRJEB13551 | SAMEA3931105 |
| ERR1359731 | England | Non-invasive | 3.93 | 2014 | 2 | | PRJEB13551 | SAMEA3930761 |
| ERR1359759 | England | Non-invasive | 3.93 | 2014 | 2 | | PRJEB13551 | SAMEA3930584 |
| ERR1359763 | England | Non-invasive | 3.93 | 2014 | 2 | | PRJEB13551 | SAMEA3930723 |
| ERR1359862 | England | Non-invasive | 3.93 | 2014 | 2 | | PRJEB13551 | SAMEA3930781 |
| 23GA0015 | New Zealand | Non-invasive | 3.93 | 2023 | 6 | Throat | PRJNA1100230 | SAMN41032505 |
| 23GA0506 | New Zealand | Invasive | 3.93 | 2023 | 6 | Blood | PRJNA1100230 | SAMN41032978 |
| 23GA0008 | New Zealand | Invasive | 3.93 | 2023 | 6 | Blood | PRJNA1100230 | SAMN41032498 |
| 23GA0256 | New Zealand | Non-invasive | 3.93 | 2023 | 6 | Throat | PRJNA1100230 | SAMN41032739 |
| 23GA0600 | New Zealand | Invasive | 3.93 | 2023 | 6 | Blood | PRJNA1100230 | SAMN41033069 |
| 23GA0688 | New Zealand | Invasive | 3.93 | 2023 | 6 | Blood | PRJNA1100230 | SAMN41033151 |
| 23GA0435 | New Zealand | Invasive | 3.93 | 2023 | 6 | Blood | PRJNA1100230 | SAMN41032912 |
| 23GA0458 | New Zealand | Invasive | 3.93 | 2023 | 6 | Blood | PRJNA1100230 | SAMN41032934 |
| 23GA0432 | New Zealand | Non-invasive | 3.93 | 2023 | 6 | Tissue | PRJNA1100230 | SAMN41032909 |
